# *RFC1* repeat expansion analysis from whole genome sequencing data simplifies screening and increases diagnostic rates

**DOI:** 10.1101/2024.02.28.24303510

**Authors:** Roisin Sullivan, Sai Chen, Christopher T Saunders, Wai Yan Yau, Yee Yen Goh, Emer O’Connor, Natalia Dominik, Valentina Galassi Deforie, Heba Morsy, Andrea Cortese, Henry Houlden, Michael A. Eberle, Jana Vandrovcova

## Abstract

Biallelic expansions and a motif change in *RFC1* are a common cause of cerebellar ataxia, neuropathy, and vestibular areflexia syndrome. Molecular diagnosis relies on a complicated combination of repeat primed PCR and Southern blotting. We developed a whole genome sequencing based method for *RFC1* repeat detection.

The combination of sequence motifs and allele length analysis in 29,478 individuals showed that 92.5% of samples have no expanded allele or have expansions of known benign motifs on one allele. In total, 103 samples were classified as biallelic carriers of pathogenic expansions and the frequency of the most common pathogenic allele AAGGG was estimated as 3.9%. Our *RFC1* classification method was validated by molecular diagnostic methods achieving sensitivity and specificity of 100% and 97.4%, respectively. Additionally, we catalogued 8 rare repeat motifs, further elucidating the high sequence complexity within the *RFC1* locus.

In this study we report a new method to identify patients carrying *RFC1* pathogenic repeat expansions from WGS data, reducing the need for the inefficient molecular workflow currently used for clinical diagnoses. This method correctly identifies recessive pathogenic repeat expansions in *RFC1* and allows screening of existing large scale WGS datasets of unsolved cases.

## Introduction

Cerebellar ataxia, neuropathy, and vestibular areflexia syndrome (CANVAS, OMIM #614575) is caused by a core triad of symptoms including cerebellar impairment, bilateral vestibular and somatosensory impairment. In 90% of cases, CANVAS is caused by a biallelic motif change together with an expansion of an intronic short tandem repeat in the replication factor C subunit 1, *RFC1*, gene [1, 2]. Since its discovery in 2019, in CANVAS and late-onset ataxia cases, the phenotypic spectrum of *RFC1* related disorders has expanded and patients often present with sensory neuropathy and/or balance control problems but without the full core triad of symptoms prompting the need to screen a wider group of patients presenting with adult-onset neurological disorders.

Whilst many short to moderate size expansions can be identified and genotyped from whole genome short read sequencing data (WGS) using bioinformatics tools such as ExpansionHunter [3, 4], the *RFC1* locus presents two challenges. The first is a large repeat size, and the second is the importance of motif sequence. As CANVAS is inherited in a recessive manner an accurate diagnosis requires quantification of both the pathogenic motif fractions and repeat length for each allele. *RFC1* repeat length can reach up to 2750 repeating units (13,750 bp) which prohibits reliable sizing of expanded alleles in WGS, but non-expanded alleles can be accurately identified. In the case of *RFC1* repeat expansions, it is also primarily the repeat motif sequence which dictates pathogenicity [5], not the allele length alone. To date, two motifs have been reported to be pathogenic, AAGGG_(n)_ and ACAGG_(n)_ [2, 6], but a large-scale screening of *RFC1* motifs and their population specificity has not been conducted yet. Current laboratory workflow developed for an accurate molecular diagnosis of *RFC1* repeat expansions involves a combination of flanking PCR and repeat-primed PCR (RP-PCR) followed by a Southern blot analysis, which is considered the gold standard for sizing repeat expansions [2, 7, 8]. In the case of *RFC1*, RP-PCR needs to be performed for each pathogenic repeat motif using sequence specific primers. A major limitation of this workflow is time-inefficiency, particularly with regards to Southern blotting, which also requires a large amount of high-quality genomic DNA. The need for motif specific primers also prevents detection of novel motifs.

We developed a method that extends ExpansionHunter and investigates repeat motifs and their fractions from WGS. Using data generated for patients recruited to the Genomics England 100,000 Genomes Project, we have shown that our method is fast and can be used at scale to investigate motif composition of the *RFC1* locus. Our classification of *RFC1* carrier status was successfully validated using gold standard molecular methods and resulted in novel diagnoses.

## Materials and Methods

### Study subjects

In total 48736 individuals recruited into the 100,000 Genome Project rare diseases cohort were included in the study (The National Genomic Research Library v5.1, Genomics England. https://doi.org/10.6084/m9.figshare.4530893.v7). Of these 29478 were probands and 19258 were relatives. About a quarter of probands (n=6557) were recruited into the neurology domain. The main disease phenotypes (n > 300) included epilepsy (n=1330), hereditary ataxia (n=886), Charcot-Marie-Tooth disease (CMT) (n=529), early onset and familial Parkinson’s disease (n=503), hereditary spastic paraplegia (n = 416), congenital myopathy (n=365) and early onset dystonia (n=308).

A cohort of 110 individuals recruited into the 100,000 Genomes Project through the National Hospital for Neurology and Neurosurgery (NHNN) were included in the validation of the predicted *RFC1* carrier status. These samples were put forward for *RFC1* screening at the NHNN due to their CANVAS-like clinical features. Genomic DNA was extracted from blood and investigated under approval of the joint ethics committee of the UCL Queen Square Institute of Neurology and the NHNN, London, UK (UCLH: 04/N034).

### Methods

ExpansionHunter, a graph based STR caller, allows the user to align reads to repeats comprised of different repeat sequence motifs [4]. The *RFC1* repeat definition in ExpansionHunter enables better alignment to both the reference (AAAAG) and the more common pathogenic repeat (AAGGG) by defining the repeat as “AARRG” where “R” represents A/G using the IUPAC degenerate base symbols. While this process is expected to improve the read alignments, other repeats, such as the rarer pathogenic motif ACAGG will also be aligned but they will be expected to have more mismatches within the graph.

Using the *RFC1* genotypes generated by ExpansionHunter v3.0 and GRCh38 reference, we first identified samples with expanded read length. Since repeat sizing can become increasingly inaccurate beyond the read length, we defined the normal, non-expanded, allele as any allele that was shorter than the read length (< 30 repeats in this study).

For samples with at least one expanded repeat, we developed a method to identify sequences of the repeat motifs within the expanded allele(s) using the following steps: (1) Extract out the part of the sequence reads aligned to the repeat section of the graph. (2) For extracted reads, collect base quality scores and an alignment segment. For each repeat in the graph extract out the aligned segment and corresponding base quality scores from the sequence read. (3) Collect and store each sequence string and fraction of high-quality bases (phred score >Q20) for each path through the repeat.(4) Combine all the read-level data for all the reads that can be associated with an expanded haplotype. For samples where both alleles were expanded, all the reads that overlap the repeat were used. For samples where only one allele is expanded, only reads that contain at least two repeats more than the number of repeats in the shorter allele were used. This ensures that the reads originate from the expanded haplotype. Using reads from expanded haplotypes, all 5-base repeat motifs observed were combined to get a complete summary of the possible repeats they contained. Due to elevated sequencing errors in repeat regions, we applied an additional filter to exclude repeat motifs that may have been present in the data due to sequencing errors. This filter required that a repeat motif was observed twice with all high-quality bases. (5) In the final step, base-quality weighted fraction of the total repeats observed in the data for known pathogenic repeats (e.g., AAGGG or ACAGG) was calculated and several heuristic rules applied to define the most likely *RFC1* repeat carrier status (Figure 1). A sample was considered ‘normal’ unless one of the following was observed: (a) The sample had biallelic pathogenic expansions if it had two expansions and >80% of the counted repeat motifs are one of the known pathogenic motifs. (b) The sample was a carrier if it had two expansions and 30-80% of the counted repeat motifs were one of the known pathogenic motifs or reads containing pure pathogenic motifs were detected (>80% pathogenic motifs within a read). Sample was also classified a carrier if it had one expanded allele and >80% of the counted repeat motifs were one of the known pathogenic motifs.

**Figure 1.**
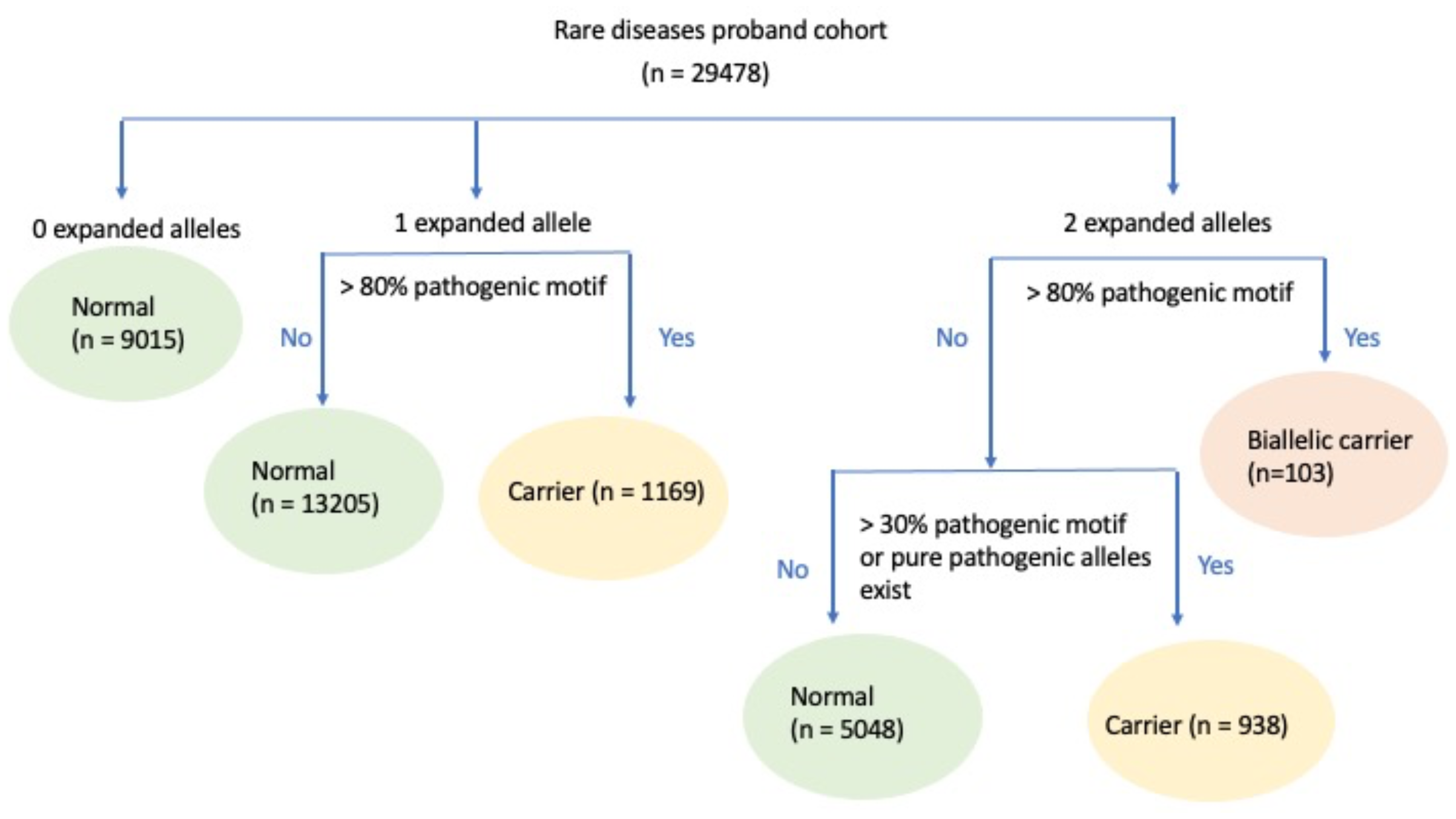
Decision tree representing algorithmic classification of *RFC1* status. ExpansionHunter repeat calls were first classified by the presence of an expansion on either allele. Individuals were classified as ‘normal’ (no expansions) if no expansions were detected on either allele. If an expansion was detected on one allele, then individuals were classified as either heterozygous carriers if the motif configuration was comprised of >80% pathogenic motifs, or normal if not. If both alleles were expanded, individuals were classified as biallelic carriers if the motif configuration comprised of >80% pathogenic motifs. If the expansions were comprised of <80% pathogenic AAGGG motifs, individuals were classified as heterozygous carriers if the expansion was comprised of >30% pathogenic motifs or alleles with pure pathogenic motif (>80% of motifs within a read) were identified.

### RFC1 screening using molecular diagnostic testing

*RFC1* molecular diagnosis was performed using a standard workflow as described previously [2]. In brief, PCR and RP-PCR were carried out, with Southern blot confirmation performed on suspected biallelic cases.

## Results

### RFC1 genotype and motif distribution

The median allele length in our cohort of 29478 rare disease probands was 12 repeats (min=5, max=144). Assuming that allele length can be reliably genotyped to a maximum of 30 repeats in 150bp sequencing data, we set our definition of normal (non-expanded) allele length as less than 30 repeats. Using this definition, 32,390 (55% of the total number of alleles) were classified as normal halving the number of alleles for further analysis. In total, 9015 (30.6%) of samples had both alleles within the normal allele length range. As expected, the majority of these samples (97.8%) contained reads which were comprised of >80% of the reference AAAAG motif.

### Analysis of samples with one expanded allele

Reference *RFC1* motif (AAAAG) was also predominant in samples with one non-expanded allele and one expanded allele. In total, we identified 14374 (49%) probands with this configuration. Motif analysis of expanded alleles identified 9158 samples with repeats composed of > 80% of reference motif. 1169 samples had >80% of AAGGG_(n)_ motifs and were classified as AAGGG_(n)_ heterozygous carriers. No ACAGG_(n)_ carrier was detected. The remaining 4047 samples had a combination of other motifs, with the majority of samples having combinations of reference, AAAGG_(n)_ and AAGAG_(n)_. Only seven samples had expanded repeats comprised of >80% of other motifs (AAACG _(n)_ (n=1), AAGGC_(n)_ (n=2), CAGGG_(n)_ (n=1), AGGGG_(n)_ (n=3)). The AAGGC_(n)_ motif was identified in two samples of Asian ancestry, AAACG _(n)_ in a proband of mixed ancestry and the rest were in probands of European ancestry.

### Analysis of samples with two expanded alleles

In samples where both alleles were expanded (n = 6089) we investigated motif fractions in all reads as well as motif purity of individual reads. Samples with >80% of AAGGG_(n)_ (n=102) or ACAGG_(n)_ (n=1) pentamers were considered to be biallelic carriers of a pathogenic motif. Apart from AAAGG_(n)_ and AAGAG_(n)_ we did not identify any other motifs present in >80% of reads. Samples with 30-80% of pathogenic AAGGG_(n)_ pentamers were classified as likely heterozygous carriers. In the remainder of samples, we analysed motif purity within individual reads to identify samples with pure >80% AAGGG_(n)_ reads and classified these samples also as heterozygous carriers of pathogenic expanded alleles. All other samples were classified as normal by our method. Fractions of common motifs within samples with two expanded alleles are shown in Figure 2.

**Figure 2.**
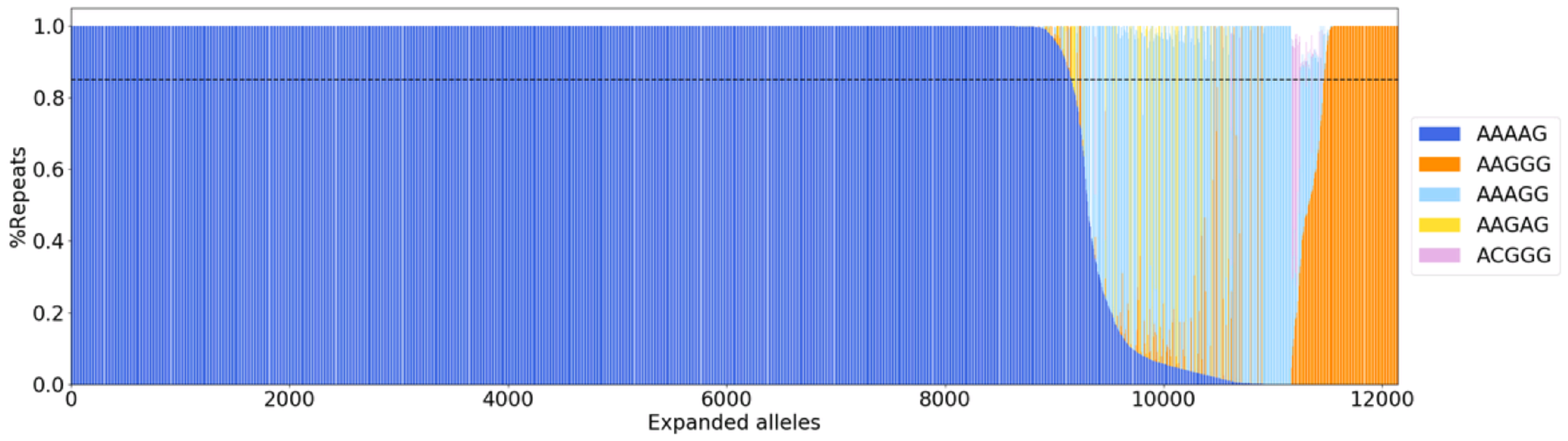
Motif distribution within the rare diseases’ cohort from the 100,000 Genomes Project data. Proportion of top five motifs within expanded alleles identified in the rare diseases’ cohort within the 100,000 Genomes Project.

Eight additional rare motifs, AACAG_(n)_, AACGG_(n)_, AAGGC_(n)_, AGGAC_(n)_, AGAGG_(n)_, AGGGG_(n)_, AGGGC_(n)_were identified and were present in both neurology and non-neurological cohorts. Three motifs (AAGGC_(n)_, AGGGC_(n)_, AGGAC_(n)_) were found in individuals with Asian or mixed Asian ancestry. In only three samples, rare motifs were observed in heterozygous carriers of pathogenic expansions. One sample had a combination of AAGGG_(n)_/AGGGC_(n)_, the second had AAGGG_(n)_/AGAGG_(n)_/AAAGG_(n)_and the final contained AAGGG_(n)_/AAGGC_(n)_repeat motifs. All three samples were from the neurology cohort.

### RFC1 classifier results

Based on our *RFC1* carrier status definition (Figure 1) the majority of probands were classified as normal (n=27,269 (92.5 %)) or as heterozygous carriers of *RFC1* pathogenic motifs (n=2107 (7.2%)). Only 103 (0.3%) probands carried two pathogenic expansions suggestive of CANVAS spectrum disease. There was no correlation between allele length calculated by Expansion Hunter and *RFC1* carrier status for expanded alleles. The overall AAGGG_(n)_ allele frequency, as defined by the number of heterozygous and homozygous carriers, was 4.6% in the neurology domain and 3.9% for the rest of the samples. Despite the presence of biallelic carriers of pathogenic expansions outside of the neurology domain, we observed a significant enrichment of biallelic pathogenic expansions in probands in the neurology cohort compared to the rest of the probands (expected (n=14) versus observed (n=69), Pearson p-value = 8.96e-53) supporting the pathogenic role of *RFC1* spectrum diseases. The AAGGG_(n)_ allele and genotype frequencies in the non-neurology cohort were in Hardy-Weinberg equilibrium.

### Phenotypic distribution of samples with RFC1 biallelic repeat expansions

The majority of biallelic carriers (n=70) were identified amongst probands recruited into the neurology domain and 33 spread across other rare disease types (paediatrics, cardiovascular, renal, hearing and sight and metabolism). The most common clinical diagnosis was hereditary ataxia, followed by Charcot-Marie-Tooth disease (n=53). In addition to these patients HPO terms ataxia and/or neuropathy were listed in 5 patients. About a third of probands (28.2%) were below the age of 54 which is the average age of neurological onset in patients with *RFC1* CANVAS spectrum disorder. Of the samples with available ethnicity details (n=102), four samples were of mixed ethnicity background and the ACAGG_(n)_ patient was of Asian ancestry. The remainder were of European ethnic background.

### RFC1 classifier validation

To confirm our results, we analysed 110 samples that were recruited to the 100,000 Genomes Project and were also put forward for molecular genetic testing of *RFC1* at the NHNN due to their CANVAS-like clinical features. Based on the molecular analysis using RP-PCR and Southern blot, 72 (64.5%) of samples were normal, nine were heterozygous AAGGG_(n)_ carriers (8.2%) and 30 (27.3%) were biallelic AAGGG_(n)_ carriers. Our method correctly classified all samples carrying the biallelic AAGGG_(n)_ repeat and eight out of nine heterozygous carriers. The incorrectly classified sample was a false negative labelled as normal, with one short and one expanded normal allele using our method but showing one pathogenic expanded allele on molecular analysis. Visual inspection of motifs within the expanded allele in this sample showed 50% of AAGGG_(n)_ pentamers, which is below our 80% threshold. The derived specificity of our classifier was 100% and sensitivity was 97.4%.

### Segregation of the AAGGG_(n)_ allele

We further assessed *RFC1* classifier reliability by segregation analysis of available trios within the rare diseases’ cohort (n=9629). In 8228 (85%) of trios all three members had no pathogenic expansion and in 33% one parent was a carrier. In 50% of these cases where one parent was a carrier, the pathogenic allele was inherited by the proband. There was no difference in paternal and paternal transmission. Of the 47 trio cases where both parents were identified as carriers of the pathogenic expansion, 13 (24%) of probands were identified as homozygotes, 29 (49%) were identified as carriers, and 18 (27%) as normal following segregation ratios expected for an autosomal recessive disease. Violations of Mendelian inheritance were identified in less than 0.3% of families; the majority of which had *RFC1* carrier proband with only normal alleles observed in parents.

## Discussion

In this study we report a new method to identify patients carrying *RFC1* pathogenic repeat expansions from WGS data. This method correctly identifies recessive pathogenic repeat expansions in *RFC1* and vastly improves carrier status detection compared to ExpansionHunter alone.

ExpansionHunter combined with WGS data from the 100,000 Genomes Project, has recently been reported as an effective approach for detecting short repeat expansions, with 97.3% sensitivity and 99.6% specificity [3]. Our method builds upon this work by demonstrating the ability to detect repeat expansions with variable motifs and has the potential to be used as a pre-screening method in a diagnostic setting. Our method correctly identified all carriers of the biallelic pathogenic repeat expansion and had a 97.4% sensitivity to detect any carrier of a pathogenic expansion. Our validation cohort was small, but the reliability of the classifier was further confirmed by segregation in available trios. Segregation analysis of the *RFC1* repeat expansion suggested a recessive mode of inheritance rather than *de novo*, with 24.5% of carrier parent trios also having a proband child identified as a patient, and 49% having a carrier proband. In trios only 0.3% of probands had no carrier parents, suggesting cases of a *de novo* variant. Upon visual inspection we identified parental samples labelled as normal to be mosaic carriers. In the present study the threshold for carrier calls was >80% of the counted repeat motifs which may need to be adjusted in the future. Large cohorts of laboratory validated samples will be necessary to fine tune the algorithm.

The pathogenic AAGGG_(n)_ allele frequency was estimated as 3.92% from the cohort of rare disease patients that were recruited outside of the neurology domain. This frequency is high for rare disease but is within the allelic frequency range published within the literature of 0.7%– 4% [2, 9-11]. Of the 103 potential patients identified, 51.5% had a clinical diagnosis of hereditary ataxia or Charcot-Marie-Tooth disease and an additional 5 probands were reported to present with ataxia or neuropathy. We also observed an enrichment of biallelic expansions in the neurology group supporting disease association between *RFC1* repeat and neurological disorders. Identification of biallelic pathogenic carriers outside of the neurology domain is likely explained by the age of probands. In our cohort 28.2% were younger than the average age of symptoms reported for CANVAS-spectrum patients and by the lack of clinical details for phenotypes unrelated to disease for which patients were recruited for.

At the time of our study only two pathogenic motifs, AAGGG_(n)_ and ACAGG_(n)_, were published in the literature, but more recent studies identified samples with rare combinations of *RFC1* repeat motifs and CANVAS-spectrum phenotype. We identified 13 motif configurations that comprised greater than 10% of motifs observed in expansions, including several motifs that have already been published, AAAAG_(n)_, AAAGG_(n)_, ACAGG_(n)_, AAGAG_(n)_ and AGAGG_(n)_, as well as 8 novel motifs [2, 6, 8], two of which have now been reported as pathogenic. As more motifs are discovered, our *RFC1* carrier decision tree can be easily adapted without the need of rerunning pentamer extraction. Detection of novel motifs in patients of non-European ancestry warrants further studies in individuals with a diverse genetic ancestry.

The method presented here can accurately detect biallelic pathogenic repeat expansions in *RFC1*, as well as other non-pathogenic motifs and is easily adaptable to other loci where motif sequence composition is of importance. It utilises outputs from ExpansionHunter and allows screening of existing large scale WGS datasets of unsolved cases which would not be feasible using current wet-lab-based molecular techniques. It also opens the possibility of a scaled down *RFC1* diagnostic workflow, which is currently a time-inefficient process.

## Acknowledgements

This research was made possible through access to the data and findings generated by the 100,000 Genomes Project. The 100,000 Genomes Project is managed by Genomics England Limited (a wholly owned company of the Department of Health and Social Care). The 100,000 Genomes Project is funded by the National Institute for Health Research and NHS England. The Wellcome Trust, Cancer Research UK and the Medical Research Council have also funded research infrastructure. The 100,000 Genomes Project uses data provided by patients and collected by the National Health Service as part of their care and support. In addition, RS is grateful to the Medical Research Council for funding this work (MRC UK MR/J004758/1, G0802760, G1001253). J.V. holds a fellowship from the Health Education England Genomics Education Programme. H.H. is grateful to the Medical Research Council (UK), the Wellcome Trust Synaptopathies Award, Ataxia UK, Rosetrees Trust, Brain Research UK, University College London Official Development Assistance and Low and Middle Income Country award, The Multiple System Atrophy Trust, Muscular Dystrophy UK, and Muscular Dystrophy Association. Andrea Cortese was supported by the Medical Research Council (MR/T001712/1), Fondazione Cariplo (grant n. 2019-1836), the Inherited Neuropathy Consortium, and Fondazione Regionale per la Ricerca Biomedica (Regione Lombardia, project ID 1751723).

## Data Availability Statement

Research on the de-identified patient data used in this publication can be carried out in the Genomics England Research Environment subject to a collaborative agreement that adheres to patient led governance. All interested readers will be able to access the data in the same manner that the authors accessed the data. For more information about accessing the data, interested readers may contact or access the relevant information on the Genomics England website: https://www.genomicsengland.co.uk/research.

